# Family Caregivers’ Experience in the Provision of Palliative Care for Adult Cancer Patients in Low and Lower-Middle-Income Countries: A Qualitative Systematic Review Protocol

**DOI:** 10.1101/2025.07.06.25330964

**Authors:** H.G.K.T. Karunarathna, V.R. Uduwela, G. Godage, N.A.Y. Isuruni, K.L.M.D. Seneviwickrama

**Affiliations:** Ministry of Health. Sri Lanka; University of Peradeniya. Sri Lanka; Fulbright Research Scholar. United States of America; Centre for Cancer Research, University of Sri Jayewardenepura, Nugegoda, Sri Lanka, Department of Community Medicine, University of Sri Jayewardenepura, Faculty of Medical Sciences, Nugegoda, Sri Lanka

**Author notes:** Corresponding Author: Prof K.L.M.D. Seneviwickrama, Centre for Cancer Research, University of Sri Jayewardenepura, Nugegoda, Sri Lanka, Department of Community Medicine, University of Sri Jayewardenepura, Faculty of Medical Sciences, Nugegoda, Sri Lanka.

**Keywords:** Family caregivers, Experiences, Provision of palliative care, Adult cancer patients, Lower-middle income countries, Low-income countries

## Abstract

**Introduction:** In contrast to high-income countries, palliative care services are limited and largely undertaken by informal caregivers, often family members, in low- and lower-middle-income countries (LMICs). This review aims to explore the family caregiver experience in providing palliative care to adult cancer patients in LMICs.

**Methods and Analysis:** This qualitative systematic review will be conducted using a comprehensive search strategy on PubMed (MEDLINE), EMBASE, CINAHL, and PsycINFO without language or date restrictions. Open University’s CORE database will be searched for grey literature. Review questions will follow the PICo framework (population, phenomena of interest, and contexts). Qualitative or mixed-method studies on the experience of family caregivers of adult cancer patients (above 18 years) receiving palliative care in LMICs will be included. Study quality will be assessed using the Joanna Briggs Institute (JBI) critical appraisal tools. Findings will be synthesized using meta-aggregation, or with a narrative synthesis if pooling is not feasible. Review process will follow the JBI methodology for systematic reviews of qualitative evidence and will be reported according to the Preferred Reporting Items for Systematic Reviews and Meta-Analyses (PRISMA) framework.

**Ethics and Dissemination:** No ethical approval is required as the study involves secondary data analysis of published literature. Ethical principles of accurate reporting and transparency will be upheld. Findings will be published in peer-reviewed open-access journals and presented at academic conferences. Recommendations will be shared with policymakers and healthcare organizations responsible for the provision of palliative care for cancer patients in LMICs.

**PROSPERO registration number:** CRD420251010556

**Article Summary:** *Strengths and Limitations of this study:*

- Four databases, as well as grey literature, will be searched using the extensive search strategy by a multidisciplinary team with expertise in systematic reviews and cancer care in LMICs.
- A rigorous methodology will be adhered to throughout the study as per the JBI guidelines for qualitative systematic reviews.
- While language was not limited in the search, due to translation constraints, only selected non-English articles will be reviewed. This limitation may introduce language bias through the exclusion of some non-English qualitative study articles.
- Due to the limited number of qualitative studies on cancer palliative care in LMICs and possible low quality of available studies, the certainty of evidence generated through this systematic review may be limited.

## 1. Introduction

Palliative care is a specialized approach aimed at improving the quality of life of patients and their families facing life-threatening disease conditions. Palliative care helps prevent and ease suffering through early identification, accurate assessment, and effective treatment of pain and other physical, psychosocial, or spiritual challenges.^1^

Cancer palliative care is a holistic approach aimed at improving the quality of life for patients with advanced cancer by addressing physical symptoms, psychological distress, and spiritual concerns. This multidisciplinary care focuses on alleviating pain, managing symptoms, and providing emotional support to both patients and their families, ensuring comfort and dignity throughout the disease trajectory.^2^

A caregiver is a person who provides services to people in need of help to take care of themselves. Caregivers may include healthcare professionals, social service workers, family members, close companions, or spiritual advisors, offering care either at home or in a healthcare setting. Caregiving can be burdensome, leading to significant physical, emotional, and financial strain, which in turn leads to negative consequences on the patients they provide care ^3 5^;. Qualitative systematic reviews have identified the needs of cancer caregivers, including adequate support, access to information, respite care, financial aid, and emotional support.^6^

A family caregiver refers to an unpaid individual, often a relative, who provides physical, emotional, and psychological support to a loved one with a serious illness. They usually have not received any formal training to provide care services and are hence known as informal caregivers.^7^ Their role is pivotal in providing comfort, managing symptoms, and ensuring the patient’s dignity. Family caregivers play a critical role in the provision of palliative care, particularly in low and lower-middle-income countries (LMICs) where formal palliative healthcare services are limited and expensive.^8^

Palliative care services in high-income countries are often well-integrated into healthcare systems, with specialized teams and structured support systems. In contrast, LMICs struggle with inadequate health care policies, lack of funding, and cultural barriers that hinder the provision of palliative care.^9–10^ Therefore, understanding family caregivers’ needs and experiences is essential in addressing challenges in palliative care to improve the quality of life of the patients and their families.^11^ A qualitative approach is suggested for exploring experiences and perspectives that are difficult to quantify. It enables participants to express their thoughts, emotions, and experiences in their own words, offering a deeper and more comprehensive understanding of the phenomenon under investigation.^12^

A growing body of qualitative systematic reviews explores family caregivers’ experiences in the provision of palliative care for adult patients with cancer, highlighting common themes of emotional burden, lack of support, communication needs, and resilience. However, all these systematic reviews are based on primary studies conducted in upper-middle-income or high-income countries.^13–18^ The proposed systematic review will synthesize findings from existing primary qualitative studies based on LMICs to provide a comprehensive understanding of the experiences of family caregivers in the provision of cancer palliative care for adult patients.

The findings of this review are significant at multiple levels. At the individual level, they can help develop interventions to enhance caregivers’ well-being and coping strategies. At the community level, strengthening community-based palliative care services can alleviate caregivers’ burdens. At the policy level, the insights can inform the creation of supportive policies, including financial assistance, training programs, and mental health support for family caregivers.

## 2. Review Question

This systematic review aims to answer the following question: “ What are the experiences of family caregivers providing palliative care for adult cancer patients in LMICs?”

This question is framed using the PICo mnemonic:

⍰ Population (P): Family caregivers (informal, unpaid caregivers such as relatives, spouses, or children).
⍰ Phenomenon of Interest (I): The experience of caregiving in palliative care settings for adult cancer patients.
⍰ Context (Co): Low- and lower-middle-income countries (LMICs), where access to formal palliative care services is often limited.

## 3. Inclusion Criteria

Inclusion criteria were developed based on PICo elements: Population, Phenomena of Interest, and Context.

### 3.1 Participants

Family caregivers who provide or have provided palliative care for adult (more than 18 years of age) cancer patients will be considered as participants in this review. Family caregivers will include parents, spouses, children, siblings, or other relatives who play a major role in the provision of palliative care for the cancer patient. The WHO definition of palliative care will be used in the identification of studies suitable for the review.

### 3.2 Phenomena of Interest

This review will consider studies that explore common themes related to caregiving experiences, including but not limited to challenges related to healthcare services, financial burdens, and social support, coping mechanisms, emotional, physical, and psychological impact of caregiving, expectations, and perceived support needs.

### 3.3 Context

Studies conducted in all types of settings in LMICs, including healthcare facilities and patients’ homes, will be selected for this review. The World Bank classification at the time of publication of the study will be used to determine the income level of the country.

### 3.4 Type of studies

This review will consider qualitative studies exploring family caregivers’ experiences in palliative cancer care, including, but not limited to, designs such as grounded theory, phenomenology, ethnography, action research, and qualitative descriptive. In addition, the qualitative component of mixed-method studies will be considered for inclusion.

## 4. Methods

The proposed systematic review will follow the Joanna Briggs Institute (JBI) methodology for systematic reviews of qualitative evidence and will be reported according to the Preferred Reporting Items for Systematic Reviews and Meta-Analyses (PRISMA) framework.^19^ The review has been registered with PROSPERO (CRD 420251010556).

### Patient and public involvement

Neither patients nor the public were involved in this protocol preparation.

### 4.1 Search Strategy

This qualitative systematic review will be conducted by the Preferred Reporting Items for Systematic Reviews and Meta-Analyses (PRISMA).^20^ A preliminary search was done in order to tailor specific search terms. Once the strategy is finalized, a comprehensive search will be conducted across PubMed (MEDLINE), EMBASE, CINAHL, and PsycINFO databases. Studies published from the inception of the database to May 2025 will be considered for this review. The detailed search strategy for each database is attached in Supplementary File 1.

The proposed search strategy for MEDLINE is given below as per the different domains considered which were combined by the Boolean operator ‘AND’:

#### 1. Family Caregiver Terminology

“ Family caregiver” OR “ informal caregiver” OR “ family member” OR “ caregiving relative” OR “ carer” OR “ spouse caregiver” OR “ parent caregiver”

#### 2. Experience Terminology

“ Experience” OR “ perception” OR “ perspective” OR “ burden” OR “ challenges” OR “ coping” OR “ expectation” OR “ needs” OR “ support needs” OR “ quality of life” OR “ psychosocial impact” OR “ mental health” OR “ burnout” OR “ psychological”

#### 3. Palliative Care Terminology

“ Palliative care” OR “ end-of-life care” OR “ terminal care” OR “ supportive care” OR “ advanced cancer care” OR “ home-based palliative care”

#### 4. Cancer Terminology

“ Cancer” OR “ neoplasm” OR “ oncology patient” OR “ malignancy” OR “ advanced cancer” OR “ metastatic cancer” OR “ terminal cancer”

In addition to database searches, the reference lists of all retrieved studies will be manually reviewed to identify further relevant articles for inclusion. Furthermore, our search strategy will also include grey literature relevant to the subject of this review. First, the CORE database by The Open University will be searched, and. second, advanced Google searches will be conducted on relevant key terms with results limited to PDF files and .edu, .gov, or .org website domains. The first 5 pages of each search will be screened for potentially relevant documents to this study.

The search strategy will use two filters, adult and human, to restrict the search to be more focused on the study population. At the initial stage, the search will include all the qualitative studies published in peer-reviewed journals related to the topic without any language filters to obtain an idea of the breadth of published literature in different languages. However, due to resource constraints, only the studies published in the English language will be included in the review. The authors will re-run the search strategy three months before the submission of the manuscript to keep the review updated.

### 4.2 Study Selection

All collected citations will be stored in the reference management software EndNote. Following the removal of duplicate records, the titles and abstracts of studies identified in the initial search will be independently screened by two reviewers using predefined inclusion/exclusion criteria to determine their relevance. This screening will be conducted on the Rayyan web application 21. The initial screening process will be broad, aiming to capture the full spectrum of studies that align with the eligibility criteria, thereby ensuring a comprehensive overview of the literature on family caregiver experiences in palliative care for cancer patients in LMICs. Discrepancies will be resolved through discussion. In case of disputes, a third reviewer will be consulted for arbitration.

Full-text versions of studies that appear to meet the eligibility criteria will be obtained and assessed for relevance by two independent reviewers. Any discrepancies regarding the inclusion of studies will be resolved through discussion and consensus between the reviewers. In cases where consensus cannot be reached, a third reviewer will be consulted to resolve the disagreement and determine final eligibility.

### 4.3 Assessment of Methodological Quality

The methodological quality of the included studies will be assessed using the JBI Critical Appraisal Checklist for Qualitative Research, which includes 10 items to evaluate research methodology and other domains of reporting.^19^ This tool evaluates key aspects such as study design, transparency in reporting, risk of bias, and reliability of findings. The critical appraisal process will adhere to the approach outlined in the published JBI systematic reviews of qualitative evidence guidelines to ensure methodological rigour and consistency.

Two independent reviewers will conduct the critical appraisal. Discrepancies in quality assessment will be resolved through discussion, and if necessary, a third reviewer will be consulted. Additional information will be sought from study investigators if required information is unclear or unavailable in the study publications/reports. The final quality assessment results will not influence study inclusion but will be considered in the synthesis and interpretation of findings. The evaluation results will be judged by the number of items that meet the standard requirement. Critical appraisal results will be reported in narrative form, and all studies, irrespective of the rating, will be included in data extraction and synthesis. A rating of 6 or lower will be considered weak (C), 7 to 8 will be considered medium (B), and 9 to 10 will be considered strong (A). Risk of bias due to missing results will not be assessed, and certainty of findings will not be assessed.

### 4.4 Data Extraction

Data will be extracted by two independent reviewers to ensure consistency and accuracy. The data extracted will include detailed information about the author, year of publication, aims/objectives (s), qualitative methods used, the population, the phenomenon of interest, the context, method of analysis, results/findings, and comments. Additionally, the specific themes related to caregiving, such as emotional burden, physical and psychological challenges, coping strategies, and social support, will be systematically extracted by the two reviewers. The extracted data will be entered into an extraction sheet, which has been developed and pretested by the review team.

The conclusions from the studies will be extracted verbatim and carefully assessed through a thorough reading and re-reading of the studies. Any discrepancies between the two reviewers during data extraction will be resolved through discussion or, if necessary, by consulting a third reviewer to achieve consensus.

### 4.5 Data Synthesis

The data synthesis will follow a meta-aggregation approach using findings from the studies that were screened and categorized. The synthesis process will involve the following :

#### Aggregation of Findings

The extracted qualitative findings will be systematically organized and categorized into distinct groups based on shared meanings. These categories will reflect various aspects of the caregiving experience in palliative care settings, including but not limited to emotional strain, coping mechanisms, and social support. Rayyan’s custom tagging and note features will be utilized in this categorization process for efficient classification and data retrieval.

#### Synthesis of Categories

The categorized findings will be consolidated into a comprehensive set of synthesized statements. These synthesized insights will enhance the understanding of the experiences of family caregivers of adult patients receiving cancer palliative care in LMICs and provide a foundation for evidence-based practice. Only unequivocal and credible findings will be included in the synthesis. Findings that are unsupported or ambiguous will be excluded from the synthesis and presented separately.

#### Narrative Synthesis

If textual pooling is not feasible, findings will be presented narratively in a detailed and rich description of caregivers’ experiences. This approach will allow for a deeper exploration of the key themes, contextual factors, and insights from individual studies.

We used the PRISMA-P checklist when writing our report.^20^

## 5. Ethical Considerations

As this study involves secondary data analysis, no ethical approval is required. However, ethical principles of accurate reporting and transparency will be upheld.

## 6. Dissemination Plan

Findings will be published in an open-access peer-reviewed journal to facilitate access to the relevant stakeholders at LMICs and will be presented at academic conferences. Recommendations will be shared with policymakers and healthcare organizations responsible for the provision of cancer care services.

## Data Availability

Data will be made available upon reasonable request from the corresponding author.

## 8. Acknowledgements

Contributions from colleagues, the University of Sri Jayewardenepura and experts who have assisted in the design, execution, or review of the systematic review will be acknowledged.

## 9. Author Contributions

MS, KK, and YI conceptualized the study. GG, VU, and MS prepared the search strategy. GG, VU, YI, KK, and MS contributed to the design of the study protocol. All authors substantially contributed to drafting and revision of the manuscript and approved the final version. All authors take responsibility for the integrity of the work as a whole. MS is the guarantor.

## 11. PROSPERO registration number

CRD420251010556

## 12. Data Statement

Data will be made available upon reasonable request from the corresponding author.

## 13. Timeline

Start date: 30 April 2025. End date: 31 December 2025.

